# A rapid review of available evidence on the serial interval and generation time of COVID-19

**DOI:** 10.1101/2020.05.08.20095075

**Authors:** John Griffin, Áine B. Collins, Kevin Hunt, Miriam Casey, David Mc Evoy, Andrew W. Byrne, Conor G. McAloon, Ann Barber, Elizabeth Ann Lane, Simon J. More

## Abstract

**Background:** The serial interval is the time between symptom onsets in an infector-infectee pair. The generation time, also known as the generation interval, is the time between infection events in an infector-infectee pair. The serial interval and the generation time are key parameters for assessing the dynamics of a disease. A number of scientific papers reported information pertaining to the serial interval and/or generation time for COVID-19.

**Objectives:** Conduct a rapid review of available evidence to advise on appropriate parameter values for serial interval and generation time in national COVID-19 transmission models for Ireland and on methodological issues relating to those parameters.

**Methods:** A review of scientific literature was conducted covering the period between December 1, 2019 and April 27, 2020. Nineteen scientific papers were evaluated in detail from 27 papers that contained information on the serial interval and/or generation time for COVID-19.

**Results:** The mean of the serial interval ranged from 3.1 to 7.5 days, based on 22 estimates, and the median from 1.9 to 6.0 days (based on 7 estimates). Only three estimates were provided for the mean of the generation time. These ranged from 3.9 to 5.2 days. One estimate of 5.0 days was provided for the median of the generation time.

**Discussion:** The values of the estimates for serial interval and generation time are heavily influenced by the contact rates between infectious and susceptible individuals. Mitigation measures that are introduced in a country or region are of paramount importance in this regard. The serial interval estimate of 6.6 days (95% confidence interval: 0.7 – 19.0) from the paper by Cereda et al.[10] is likely to be the most relevant to European countries. National estimates should be obtained as soon as possible.

**Strengths and limitations of this study:**

- The study provides timely information on serial interval and generation time for those involved in the development of models and in the implementation of control measures against COVID-19.
- This is a rapid review of available evidence in the scientific literature between December 1, 2019 and April 27, 2020 on the serial interval and/or the generation time and it contains the usual limitations associated with such a review.
- Eleven of the 19 papers reviewed in detail were pre-print articles.
- The statistical methods used in the different papers were not analysed in detail.

## Introduction

In response to the coronavirus (COVID-19) outbreak, the Irish Epidemiological Modelling Advisory Group (IEMAG) for COVID-19 was established to assist the Irish National Public Health Emergency Team (NPHET) in their decision-making during the pandemic. A subcommittee from IEMAG was tasked with researching the various parameters, leading to the development of a series of synthesis documents relevant to the parameterisation of a COVID-19 transmission model for Ireland.

The serial interval is the time between symptom onsets in an infector-infectee pair, i.e. the interval between the onset of symptoms in an infectee and its presumed infector. This can be a negative number if the onset of symptoms in the infectee occurs prior to the onset of symptoms in the infector. The generation time, also known as the generation interval, is the time between infection events in an infector-infectee pair. The serial interval and the generation time are key parameters for assessing the dynamics of a disease. The generation time or its proxy, the serial interval, is an essential quantity for determining the reproduction number.

A number of scientific papers reported information pertaining to the serial interval and/or generation time for COVID-19. In the context of national control efforts in Ireland, our objective was to conduct a rapid review of available evidence to advise the IEMAG on the appropriate parameter values for serial interval or generation time in national COVID-19 transmission models and on methodological issues relating to those parameters. This information may also be of use to developers of models and those involved in the implementation of control programmes in other countries.

## Material and methods

The guidelines in the protocol “Rapid reviews to strengthen health policy and systems: A practical guide” produced by the World Health Organization were used for carrying out this review. This can be accessed at https://apps.who.int/iris/bitstream/handle/10665/258698/9789241512763-eng.pdf;jsessionid=E033D9A6E3118CE0701D03815D63F648?sequence=1. The PRISMA-ScR checklist (https://www.equator-network.org/wp-content/uploads/2018/09/PRISMA-ScR-Fillable-Checklist-1.pdf) for scoping reviews was also used.

We conducted a review of the literature between December 1, 2019 and April 27, 2020 for all countries. Publications on the electronic databases PubMed, Google Scholar, MedRxiv and BioRxiv were searched with the following keywords: “Novel coronavirus” OR “SARS-CoV-2” OR “2019-nCoV” OR “COVID-19” AND “serial interval” OR “generation time” OR “generation interval”. In view of the fact that very limitied information was likely to be available on serial interval and generation time for COVID-19, all relevant publications, including pre-print papers, were considered for possible inclusion. Bibliographies within these publications were also searched for additional resources and a manual search was also carried out. Summaries, citations and extracted parameters from these publications were added to a specifically designed database. The review was confined to papers that were published in the English language.

Papers that did not contain original parameter estimates of serial interval or generation time parameters were discarded. Some of the papers contained parameter estimates derived only from original data without the fitting of statistical distributions. As these papers were considered to be less useful for model development than papers containing distribution-derived estimates, they were also omitted from further consideration. Finally, those papers that did not provide a clear methodology on how the parameter estimate were obtained were also omitted.

Parameter estimates for the serial interval and the generation time, including means, medians and 95% confidence intervals, were extracted from the remaining papers. A critical appraisal was carried out on the retained papers with a view to identifying the most relevant findings, the strengths and weaknesses of each study and particularly the potential for bias.

Based on the parameters reported in the papers, we made simulations (n=10,000 samples) from serial interval or generation time distributions, we generated simulations (n= 10,000 samples) based on the reported distributions and generated box-plots with the aim of allowing easy comparison between the estimated distributions from different studies. The box-plots were generated using the “ggplot2” package in the R statistical environment.

## Results

Twenty seven scientific papers provided parameter estimates for the serial interval and/or the generation time. Seven of these papers[1-7] were removed as they only contained estimates derived from the original data or the methods for estimating the parameters were not clear. The paper by Zhang[8] was removed because that study used the same data as had previously been used by Du et al. [9] in estimating the serial interval.

Following the removal of these papers, 19 papers were further evaluated. The estimates for the serial interval and or generation time can be found in Table 1.

**Table 1.**
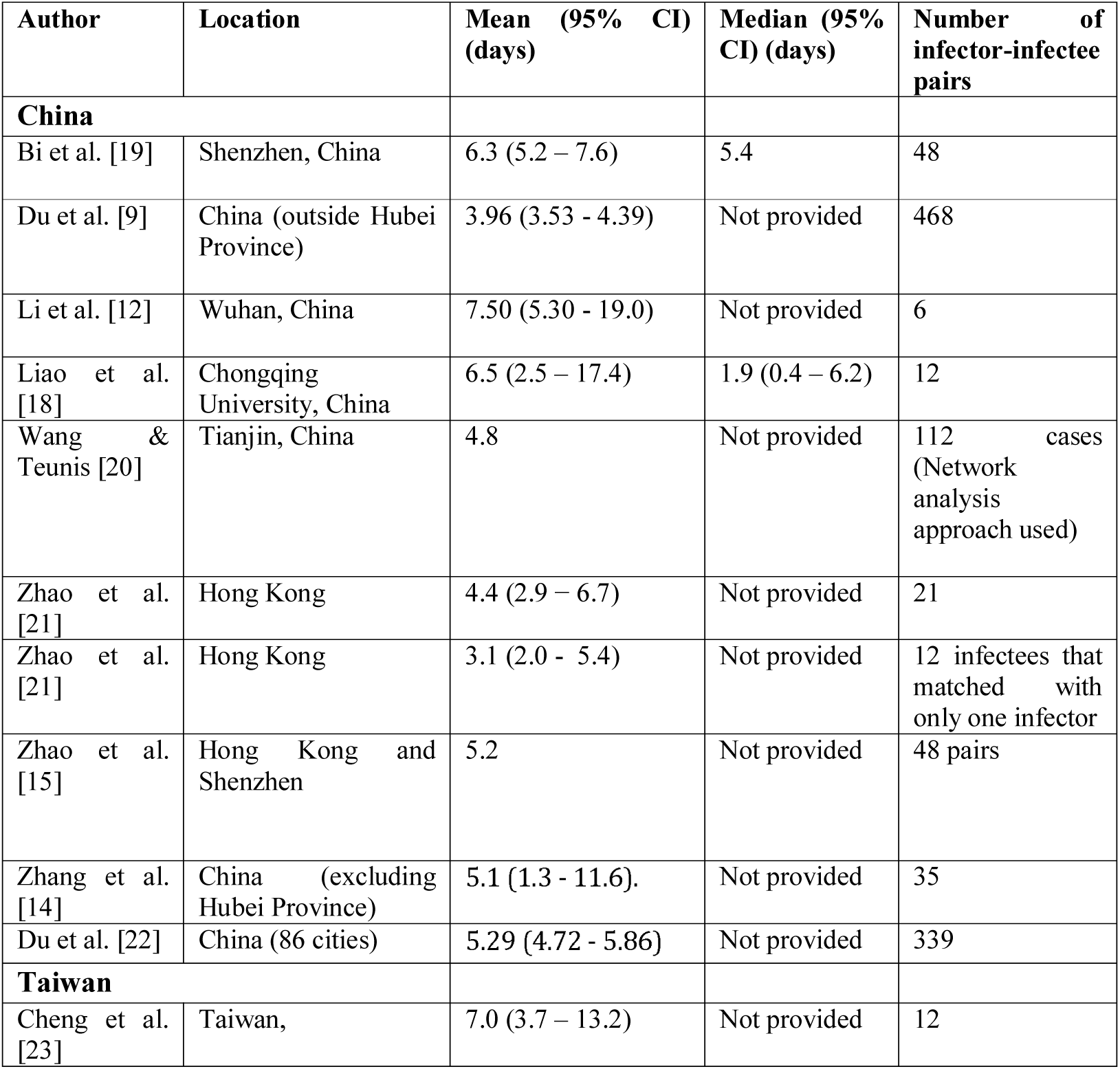
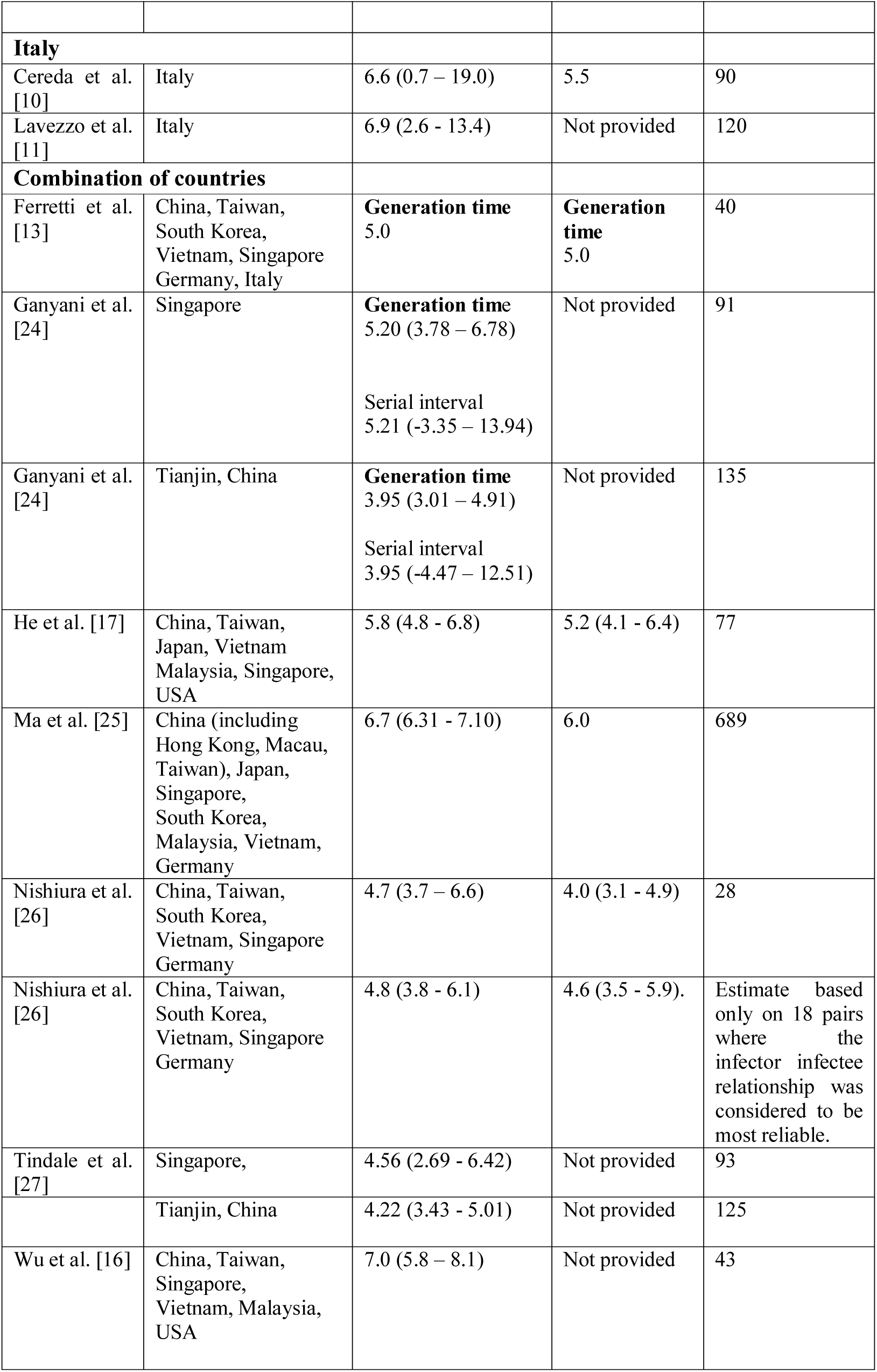
Estimates of serial interval and generation times for COVID-19 from 19 scientific papers by country. The parameter estimates relate to the serial interval unless otherwise stated.

Most of the studies relate to Asian countries, particularly China. Some of the studies also used data from Germany, Italy and the USA. However, apart from the studies by Cereda et al.[10] and Lavezzo et al. [11] which deals solely with the COVID-19 outbreak in Italy, the number of datapoints from the non-Asian countries is very small.

Eleven of the papers had a pre-print status. The published studies consisted of research articles[12-14, 19], letters[15-16,9] and a brief communication[17].

All except two studies provided estimates for the population as a whole. The study by Liao et al.[18] provided estimates for adolescents and young adults. In the study by Zhao *et al*.[15], an estimate was provided for males as well as for the population as a whole. Some of the studies provided more than one estimate.

A total of 22 estimates were provided for the mean of the serial interval. These ranged from 3.1 to 7.5 days. A total of 7 estimates were provided for the median of the serial interval. These ranged from 1.9 to 6.0 days.

Three estimates were provided for the mean of the generation time. These ranged from 3.9 to 5.2 days. One estimate of 5.0 days was provided for the median of the generation time.

A variety of statistical distributions were fitted to the data in order to get the parameter estimates. These included, normal, lognormal, gamma and Weibull distributions. Figure 1 shows boxplots of the samples simulated from these distributions (n = 24 estimates from 18 publications). Estimates were plotted from all but the study by Liao et al.[18] included in Table 1. Pending clarification from the authors, we could not replicate the distribution described by Liao er al. [18].

**Figure 1:**
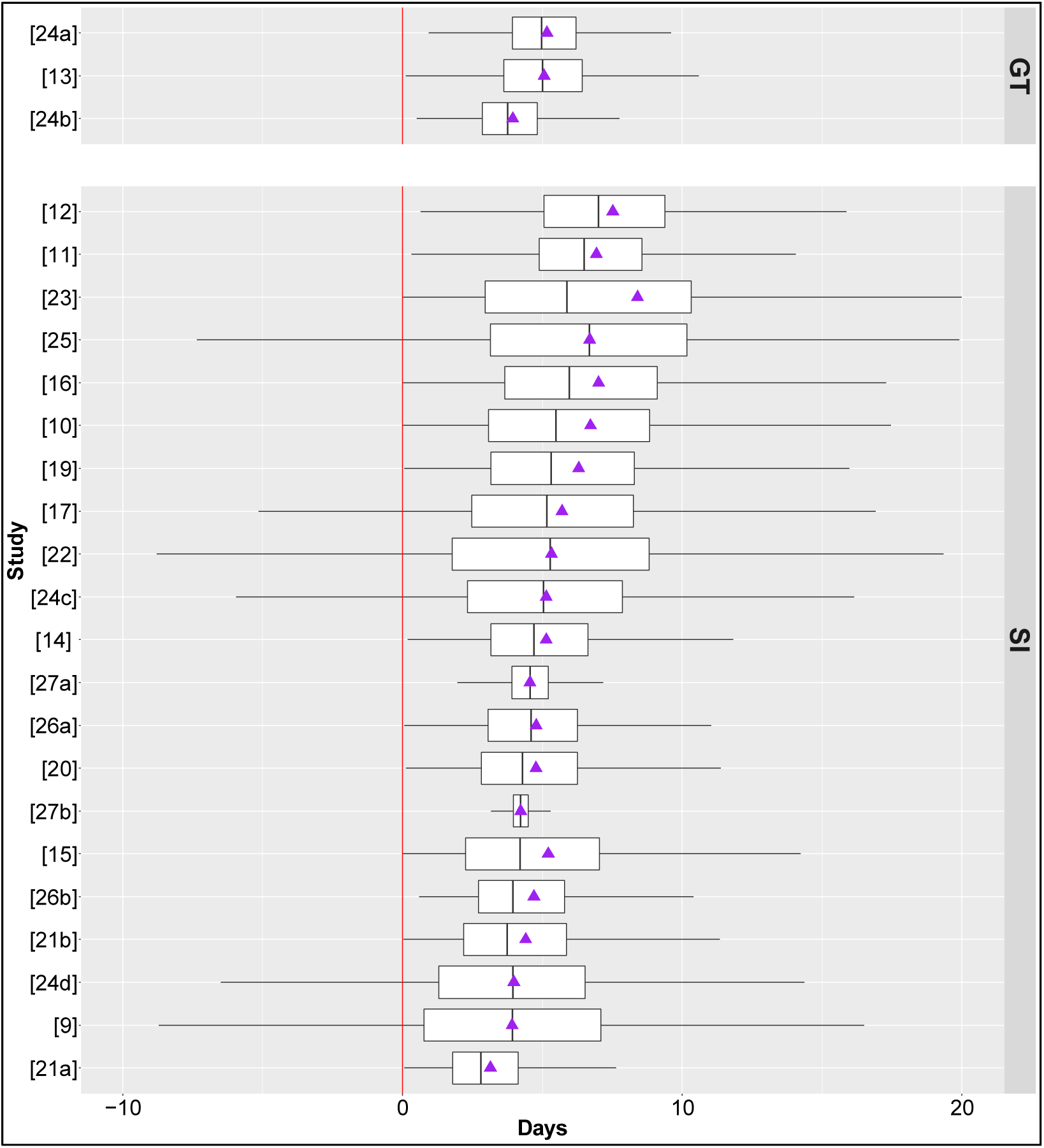
Box-plots summarising simulations (n=10,000 samples) of generation time (GT, upper plot n=3 estimates from 2 publications) or serial interval (SI, lower plot n =21 estimates from 17 publications). The purple triangles correspond to the simulated sample means. [21a]=smaller sample in study [21] of 12 more certain pairs, [21b]= sample of 21 pairs in study [21], [24a] = generation time based on Singapore data in study [24], 24b = generation time based on Tianjin data in study [24], 24c = serial interval based on Singapore data in study [24], 24d = serial interval based on Tianjin data in study [24]. 26a=smaller sample of 18 more certain pairs in study [26], 26b= sample of 28 pairs in study [26], [27a] = Singapore in study [27], 27b = Tianjin in study [27].

## Discussion

Our scientific understanding of novel emerging pathogens is dynamic and constantly evolving as new information emerges. Early estimates of key parameters are vital in assessing the natural history of a novel emerging infectious disease such as COVID-19 and the likely impact of control measures. Pre-print papers are a valuable source of information in this regard with the proviso that the quality of these will be unclear given the lack of peer review. All the studies reviewed here were also compromised by constraints that are present at the beginning of a new disease, including the lack of specific surveillance systems, information gathering systems and precise case definitions.

### Range of estimates obtained

The papers reviewed provide initial parameter estimates for the serial interval and/or the generation time for COVID-19. Most of the estimates were for the serial interval rather than the generation time because in real life infection times are rarely available, so generation times cannot be estimated. Instead, typically, the onset of symptoms is observed. The estimates for the mean of the serial interval ranged from 3.1 to 7.5 days. There are a number of reasons why the estimates are wide ranging. The interval between symptoms in an infector-infectee pair will be strongly influenced by the level of social contact. This will vary widely between different countries and indeed within countries. The impact of mitigation measures is also likely to be a key factor. The implementation of control measures will reduce the opportunity for an infected individual to transmit the disease to a susceptible individual. Consequently, the serial interval is likely to decrease during the course of an epidemic. Zhao *et al*. [15] showed that the serial interval decreased by 6.2% per day (95% CI, 0.4-11.6%) from January 10 to February 2 in Hong Kong and Shenzhen. They attributed this to the strengthening of the public health control measures over time. Stratified results produced by Bi et al.[19] showed that if the infector was isolated less than 3 days after symptom onset, the average serial interval was 3.6 days, increasing to 8.1 days if the infector was isolated on the third day after symptom onset or later. Du et al. [9] pointed out that the time between successive cases contracts around the epidemic peak and that this may have influenced their estimates. On the other hand, in a study of the Po’ municipality of Italy, Lavezzo et al. [11] estimated that the serial interval increased from 6.90 days before the implementation of comprehensive control measures to 10.12 days after the implementation of these measures. Possible reasons for this increase are not discussed in the paper.

The value of estimating the serial interval, generation time and other key parameters at the start of an epidemic was emphasized by a number of authors. As highlighted by Bi et al.[19], the study of an emerging pathogen at the time of its introduction provides a unique opportunity to characterize its transmission and natural history. In particular, it is possible to make assumptions about when and where cases were likely infected that are more difficult when the pathogen is widespread. Furthermore, during these early phases, uninfected and asymptomatic contacts are often closely tracked, providing critical information on transmission and natural history.

### Methods used for estimating the serial interval and the generation time

The estimation of the serial interval and the generation time parameters for COVID-19 presented a number of other challenges and the potential for obtaining biased estimates, as was acknowledged by a number of authors. We identified a number of specific issues in the papers that we reviewed, including the following:

- In clustered outbreaks, which is crucial to estimating the serial interval, the order of transmission (i.e., who is infector and who is infectee) can easily be mistaken. Also, given the possibility of pre-symptomatic and asymptomatic transmission particularly as the epidemic progresses, it can be difficult to determine the source of infection with certainty. In view of this, it is important that there is a well-defined methodology for determining the serial interval/generation time. Some of the studies did not describe how the order of transmission issue was handled. In other studies, efforts were made to deal with the difficulties related to the order of transmission and the true source of infection. Nishiura et al. [26] provided separate estimates of the serial interval parameter distribution for “18 most certain pairs”. Zhao et al.[21] provided separate estimates of the serial interval parameter distribution for “infectees with only one infector”. Tindale et al.[27] used a mixture model approach for serial intervals to avoid assuming that the presumed infector is always the true infector. Ganyani et al. [24] used a Markov chain Monte Carlo (MCMC) approach for the same purpose. Ma et al. [25] made an effort to overcome this issue by setting out a clear methodology for ensuring that the order of transmission was correct.
- Generally, publicly available datasets were used in the studies under review. Zhao et al. [21] mention the fact that the lack of information in publicly available datasets makes it difficult to fully interpret the data. Also as mentioned by Du et al.[9], if the data are restricted to online reports of confirmed cases, they might be biased toward more severe cases in areas with a high-functioning healthcare and public health infrastructure. The rapid isolation of such case-patients might prevent longer serial intervals, potentially shifting the estimates downward compared with serial intervals that might be observed in an uncontrolled epidemic. In general, it is likely that less severe and asymptomatic cases are underrepresented in the datasets examined.
- In some of the studies, infector-infectee pairs from a variety of countries were used to estimate the serial interval. The number of pairs from some countries were very small. For example, in the paper by He et al. [17], of the 77 pairs used, one was from the USA, one was from Singapore, two were from Malaysia, two were from Vietnam, four were from Taiwan, 12 were from Japan and the rest were from various parts of China. These cannot be considered representative of the countries from which they were drawn. The same conclusion applies to the studies by Ma et al. [25], Ferretti et al. [13], Nishiura et al.[26] and Wu et al.[16]. In other studies, pairs were drawn from particular countries or regions during particular time periods. These may have been more representative of the population from which they were drawn. However, in some cases, e.g. Li et al. [12], the number of pairs selected was very small compared to the total number of cases included in the study, again calling into question the representativeness of the pairs used to estimate the serial interval or generation time.
- The case data, including the identity of each infector and the timing of symptom onset, was based on individual recollection of past events. Du et al. [9] highlight the fact that if recall accuracy is impeded by time or trauma, case-patients might be more likely to attribute infection to recent encounters (short serial intervals) over past encounters (longer serial intervals). Therefore, it is likely that recall bias is present in all of the studies and it is not possible to distinguish the level of bias present in the different studies.
- The number of pairs used to estimate the serial interval varied considerably. Only six pairs were used in the study by Li et al. [12]. In contrast, a total of 468 pairs were used in the study by Du et al.[9] and 689 pairs were studied by Ma et al.[25]. However, the value of increased sample size must be evaluated against the difficulty of ensuring accuracy of the infector infectee relationship as the sample size increases. Lavezzo et al. [11] indicate that 120 pairs were used in their study but there is a lack of clarity on how this number was obtained.
- Cheng et al. [23] state that 12 pairs were used in their estimation of the serial interval. Figure 1 of their paper indicates that these included three cases with asymptomatic infection. It is not clear how these cases were handled in estimating the series interval. Moreover, pairs in which the exposure date of the infectee was earlier than the date of symptom onset of the infector were excluded from the study.
- In the study by Zhang et al. [14] and in other studies, the serial interval was estimated from cases in household clusters. The authors make the point that estimations based on household clusters may be 20% shorter than the true value of the serial interval.
- Zhao et al. [21] highlighted the possibility of right truncated selection bias, i.e. the possibility of infector-infectee pairs with longer SI being under-represented in the sample due to short investigation period. To minimise this possibility, the set that last date of onset symptoms for infectees as 16 days before the end of the study investigation point.

It should be borne in mind that some of the studies may have been using the same case data in estimating the serial interval or the generation time. For example, the studies by Tindale et al.[27] and Ganyani et al.[24] were carried out in Singapore and Tianjin over the same time period. Consequently, the estimates cannot be considered to be fully independent of each other. Likewise, it is possible that there was an overlap in the case data used by Du et al. [9] and Du et al. [22] as these studies were carried out by the same group of authors.

### Statistical distributions used in estimating serial interval and generation time

In most of the studies, a gamma or Weibull distribution was fitted to the data to estimate the serial interval distribution. A problem with these distributions is that negative values of the serial interval (that is, when symptoms manifest in the infectee before the infector) cannot be included. In the study by Du et al.[9], 59 of the 468 reports indicate that the infectee had symptoms earlier than the infector. Du et al. [9] and Du et al.[22] cautioned against using distributions that excluded the non-positive data and making assessments and projections based on the truncated data. In their view, the normal distribution provides the best fit for the full dataset (shifted or not) and they recommended this distribution for future epidemiologic assessments. This approach was also used by Ma et al.[25] and Tindale et al.[27]. In Ma et al.[25] study, shifted lognormal, Weibull and gamma distributions were also fitted to the data for the serial interval.

### Relationship between the serial interval, generation time and the reproduction number

The generation time is used to estimate the reproduction number. Because of the difficulty in estimating the generation time, the serial interval is often used as a surrogate for the generation time. The serial interval and the generation time will have the same mean value provided that the incubation times of the infectee and infector are independent and identically distributed but their variances are expected to be different. Britton and Scalia-Tomba[28] highlighted the fact that the difference in variance between the serial and generation time can lead to biased estimates of the reproduction number. More specifically, when the serial interval distribution has a larger variance than the generation time distribution, using the serial interval as a proxy for the generation time will lead to an underestimation of the basic reproduction number. Ganyani et al.[24] provided estimates for both parameters based on data from Singapore and China and described a method for obtaining an unbiased estimate of the generation time.

## Conclusion

Overall, the availability of parameter estimates and information on the serial interval and generation time of the COVID-19 virus is very valuable at such an early stage of the pandemic. However, the estimations are very dependent on the specific factors that applied at the time that the data was collected, including the level of social contact. Consequently, the estimates may not be entirely relevant to other environments. The serial interval estimate of 6.6 days (CI 0.7 – 19.0) from the paper by Cereda et al. [10] is likely to be the most relevant to European countries. National estimates should be obtained as soon as possible. In light of the biases that could occur, the serial interval and or the generation time should be estimated from early cases and careful consideration should be given to the methodology that is used.

## Data Availability

All data is publicly available.

## Authors contributions

Studies were selected and screened initially by AC, KH and FB using search terms outlined the methodology section, with parameters identified and recorded. This was reviewed and supplemented by a manual search by JG, again with parameters identified and recorded. JG conducted the eligibility screening of shortlisted studies, extracted the data and conducted the review with input from all authors. MC generated Figure 1. SJM, MC, ABC, AB, AWB, EAL and CGM undertook interim reviews. All authors read and approved the final manuscript.

## Patient and public involvement statement

It was not appropriate or possible to involve patients or the public in the design, or conduct, or reporting, or dissemination plans of our research

